# The mortality risk factor of severe community-acquired pneumonia (SCAP) patients with Sepsis: a retrospective study

**DOI:** 10.1101/2022.05.06.22274786

**Authors:** Cong Geng, Jun Jin, Zhejun Yu

## Abstract

**Objectives:** Sepsis is one of the most common comorbidities in severe community-acquired pneumonia (SCAP) patients. We aimed to investigate the characteristics and mortality risk factors of SCAP patients hospitalized with Sepsis.

**Design:** A retrospective, single-centre study.

**Setting:** This study was conducted at a tertiary hospital in Southern China.

**Participants:** A total of 119 patients with SCAP, aged 17 years or older, were treated in the Integrated intensive care unit from 1 January 2018 to 30 December 2020.

**Interventions:** none.

**Outcome:** 180-day mortality was the primary outcome.

**Results:** 119 patients were divided into the survivors (83 patients,69.75%), and the non-survivors (36 patients,30.25%). There are more pronounced inflammatory responses and respiratory problems at the beginning of the disease in non-survivors, requiring stronger respiratory and circulatory support. The CURB-65 score was a better predictor of mortality than the PSI and APACHE2 scores, AUCs of CURB-65: OR 0.744, *p*<0.005. For the entire treatment cycle, the non-survivors had a longer duration of persistent fever, required continuous and repeated airway intervention, and a longer duration of Vasopressor support (P<0.001). SCAP with bacterial infection as the onset, or secondary bacterial infection had a poor prognosis (P=0.018). The non-survivors had more use of different types of antimicrobials (P<0.05), because of Multidrug-resistant (MDR) organisms. And have faced more antifungal treatment failures (P=0.006). The mortality risk factors were comorbid with a duration of Vasopressors support, duration of persistent fever, age, numbers of antimicrobials for MDR organisms, CURB-65 score and duration of Neuromuscular Blocking Agents (NMBAs) (OR=1.234, OR=1.158, OR=1.084, OR=6.484, OR=3.386, OR=1.505, p<0.005, respectively).

**Conclusion:** Dynamic monitoring of the duration of patients’ abnormal indicators can help predict the prognosis. Age≥65.5 years, fever duration ≥9.5 days, number of antimicrobials for MDR organisms ≥2 types, longer NMBAs and Vasopressors use, and higher CURB-65 score were mortality risk factors in SCAP-Sepsis patients.

**strengths and limitations of this study:** We evaluated dynamic monitoring of the duration of patients’ abnormal indicators can help predict the prognosis. To the best of our knowledge, a very few studies had done a dynamic monitoring of the duration of patients’ abnormal indicators in the field of SCAP with Sepsis. The retrospective nature of the study was a limitation, statistical data including respiratory support in later treatment, can be further quantified.

## INTRODUCTION

Recently, CAP has been recognized as showing a variety of disease severity, ranging from almost asymptomatic infection to fulminant systemic disease with respiratory failure and multiple organ dysfunction. [1–2] Among over 30% of cases, severe Sepsis occurred in the early stage of infection, probably implicating multiple organ systems and associated with CAP severity and mortality.[1] Respiratory infections accounted for 40% to 60% of Sepsis causes, and Mechanical Ventilation could further increase systemic inflammation in patients with Sepsis.[3] In pneumonia cases, when endothelial and epithelial cell barriers are concentrated in the lung, the lung became a target and a source of inflammation. Cell injury of the infected organism triggered a cascade reaction of cytokines and chemokines. [4–6] 9% to 16% of CAP patients were sent to the integrated Intensive Care Unit (ICU) as severe respiratory failure, severe Sepsis or Septic Shock. The mortality was up to 50% in those who were dependent on Vasopressin, and deficiency of initial antibiotic treatment was associated with poor prognosis.[7]

According to Sepsis Management Guidelines, [8] the Quick Sequential Organ Failure Assessment (qSOFA) Score is applied to make a preliminary classification for ICU patients, SOFA and Acute Physiology and Chronic Health Evaluation II (APACHE2) scores are widely used to evaluate the prognosis of patients. The acquired data for calculation is generally the worst value 24 hours after admission to ICU. It has been suggested that Mechanical Ventilation or Vasopressin support were early risk factors with poor prognostic value.[9] Nevertheless, it’s worth noting that the majority of patients underwent deterioration of diseases during hospitalization, and the condition at admission was not always the worst time throughout the disease process. As for the immune response to Sepsis, the initial immune response is hyperinflammatory, but the response rapidly progresses to hypoinflammatory. A secondary bump in the hyperimmune state can occur during the hospital course with secondary infections. However, severe Septic Shock, may not be attributable solely to an “immune system gone haywire,” but may indicate an immune system that is severely compromised and unable to eradicate pathogens.[10]

Therefore, we hold the opinion that it may be better to apply the duration of poor factors to evaluate patients’ prognosis, and the highlight of collection may be changed from a certain point to a certain period since it is unavoidable to collect vital signs of patients. In consideration of the above questions, we collected data on the admission of patients with severe pneumonia, as well as supportive data that may affect prognosis in the process of overall disease progression. In our study, the duration of abnormal vital signs and supportive treatment were analyzed to estimate their effect on patients’ prognoses.

## MATERIALS AND METHODS

### Study design

This was a single-centre retrospective analysis. Patients with the onset of CAP and conforming to Sepsis (infection +SOFA≥2) were included in the analysis,[8] with the outcome conforming to severe pneumonia. [11–12] The primary outcome was 180-day mortality. The baseline characteristics, clinical outcomes, and prognostic factors related to mortality were assessed. The written informed consent was waived due to the observational nature of the study. Ensure patients’ anonymity.

### Subjects

We retrospectively analyzed the clinical data of 119 patients with SCAP who were treated in the Integrated ICU of The First Affiliated Hospital of Soochow University (a comprehensive tertiary adult hospital) from 1 January 2018 to 30 December 2020. The observation endpoint was 180-day mortality. Our inclusion criteria were: CAP whose outcome met the diagnosis of severe pneumonia and patients who met Sepsis criteria (SOFA≥2) at the onset. Our exclusion criteria were: patients Under 17 years of age. Patients who refused invasive resuscitation. Patients who have incomplete data. Patients whose outcome of treatment is not clear. Patients with unknown or mixed infection (more than one known infection source). Patients with Long-term Sanatoriums and Tend and Protect Hospitals treatment experience. Patients with transplantation, primary lung tumor, or advanced tumors at other sites. Presence of leukopenia or neutropenia (unless due to pneumonia). Adjuvant therapy for severe immunosuppression in human immunodeficiency virus-positive (HIV) patients (CD4 <100). Patients with previous underlying pulmonary diseases (e.g., COPD, asthma, etc.) require long-term home oxygen therapy. Figure 1 shows the flow diagram of the study.

**Figure 1.**
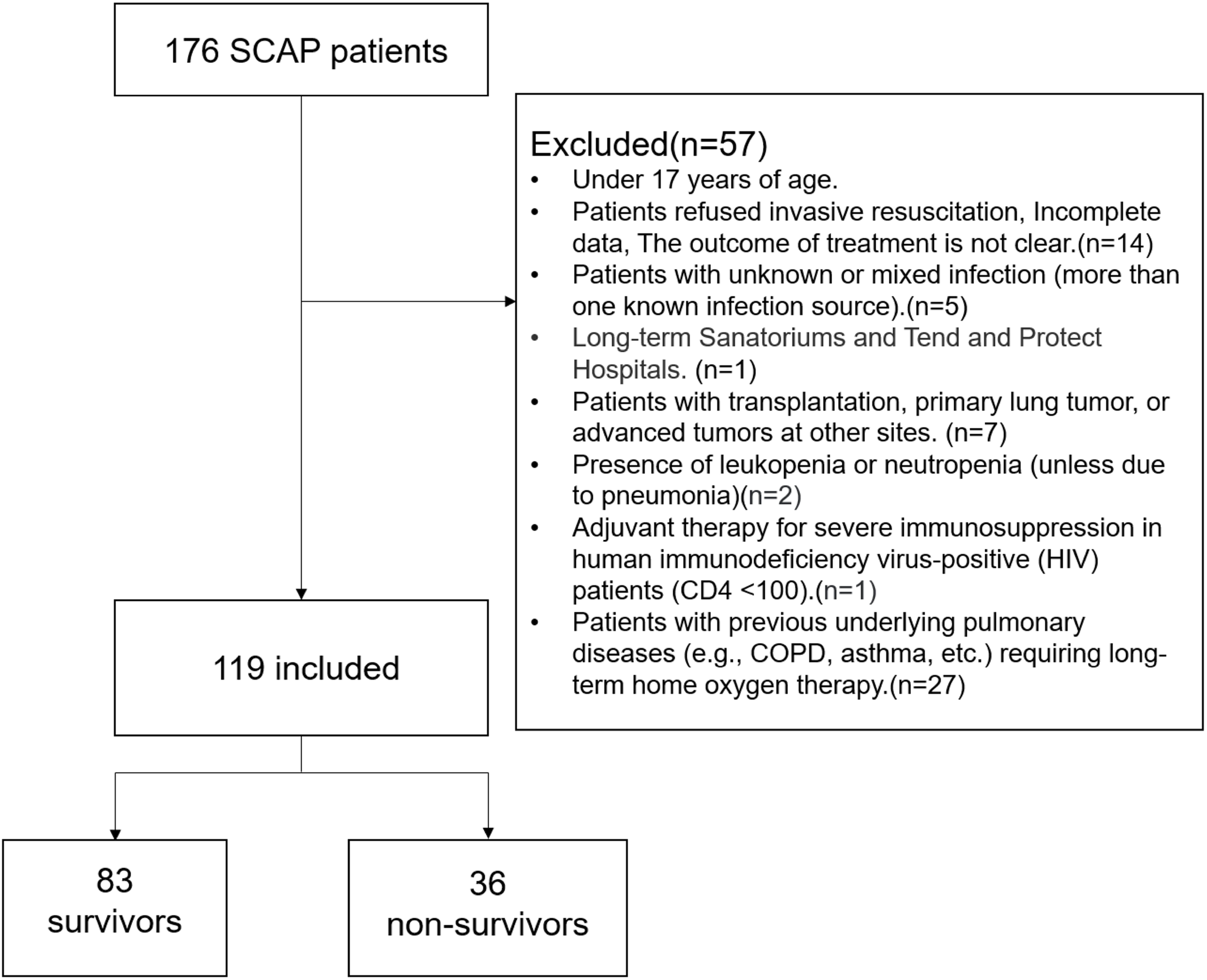
Flow chart of the study procedure. SCAP, severe community-acquired pneumonia. Invasive resuscitation includes endotracheal intubation and cardiopulmonary resuscitation.

### Data collection and definition

Our study used an electronic medical record system to collect data for retrospective analysis. Our data were recorded by attending nurses and doctors at the time of patients’ presentation to the emergency department (ED). The demographic characteristics of each patient including comorbidities were reviewed thoroughly. We collected the patients’ worst vital signs, laboratory results, ventilator support, and use of pressors within 24 hours of admission before initiation of ICU treatment. Score scales were used to calculate the relevant parameters of patients at admission, including APACHE2, SOFA, Pneumonia Severity Index (PSI), and CURB-65(a 5-point score based on confusion, urea, respiratory rate, blood pressure, and age ≥65). [13–16] (table2) Duration of fever, infection markers, respiratory management, and Sepsis medication were recorded during ICU treatment. Pathogenic microorganisms and subsequent antibiotic use of CAP were also collected.

### Microbiological Analysis and Diagnostic criteria

Microbiologic diagnostic criteria were the following:

1) Isolation of microorganisms in BAL (≥10^4^UFC/mL), BAS (≥10^5^UFC/mL) or in pleural fluid; 2) Isolation of one predominant microorganism in sputum or L pneumophila in buffered charcoal yeast extract (BCYE) agar; 3) Microorganisms in blood culture; 4) Seroconversion or a fourfold antibody increase in titers of IgG for C pneumoniae (≥1:512), M pneumoniae and C burnetii, (≥1:160) or IgM ≥1:32 for C pneumoniae, and≥1:80 for M Pneumoniae and C burnetii; 5) Positive detection of viral nucleic acids in nasopharyngeal swab. 6)Pathogens were monitored by Next Generation Sequencing. Mixed etiology was defined as pneumonia due to more than one pathogen (virus, bacteria or fungi). [17]

### Statistical analysis

Kolmogorov-Smirnov test was used to test the normality of continuous variables. The continuous variables conforming to the normal distribution were compared using the independent sample T-test and were expressed as mean (SD). All continuous variables not conforming to normality were expressed as median (IQR) and were compared by Mann Whitney test. The categorical variables were expressed as frequency and percentage and were compared using the probability ratio χ2 test. The logistic regression model was used to calculate the Odds ratio (OR) of death variables. The area under the curve (AUC) was analyzed and calculated by the receiver operating characteristic curve (ROC curve). Statistical analysis and graphic rendering were performed using SPSS26.0 and GraphPad Prism 9.0. Double-tailed p<0.05 was considered statistically significant.

## RESULTS

### Study population

Figure 1 is the flow chart of the study. A total of 176 patients were enrolled in the initial study, aged 17 years or older. Among them, 14 patients had missing data, 5 patients had multiple site infections, 1 patient lived in a nursing home for a long time, 7 patients had an advanced tumor or primary lung cancer or had undergone transplantation, and 2 patients had leukopenia or neutropenia prior to onset. 1 patient had HIV and severe immunodeficiency. 27 Patients with previous underlying pulmonary diseases (e.g., COPD, asthma, etc.) require long-term home oxygen therapy. The final number of patients included in the study was 119. At the endpoint of 180-day, 36 patients died and 83 survived. The percentage of deaths was 30.25%.

### Patient Characteristics

Table 1 shows the baseline characteristics and complications of the two groups. The non-survivors had older onset age (*p*<0.001) and was more emaciated (*p*=0.002). They had similar gender profiles and length of hospital stay (LOS). For comparison of underlying diseases, the non-survivors had a higher percentage of nervous system diseases (including cerebrovascular accidents, epilepsy, brain trauma, etc.) (*p*=0.015) and long-term Immunosuppression therapy (*p*=0.048) than the survivors. In terms of complications, except liver and deep venous thrombosis (DVT), multiple organ dysfunction syndrome (MODS) (*p*<0.001), heart(*p*=0.041), kidney(*p*=0.001), gastrointestinal tract (GIT) (*p*=0.048), nervous system(*p*=0.029), myelosuppression(*p*=0.003), and coagulation disorders (*p*=0.014) had a higher percentage in the non-survivors.

**Table1.**
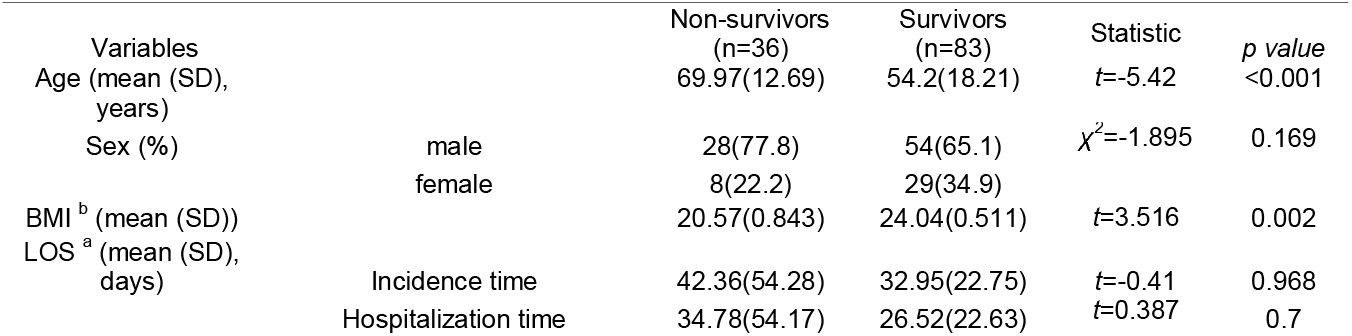
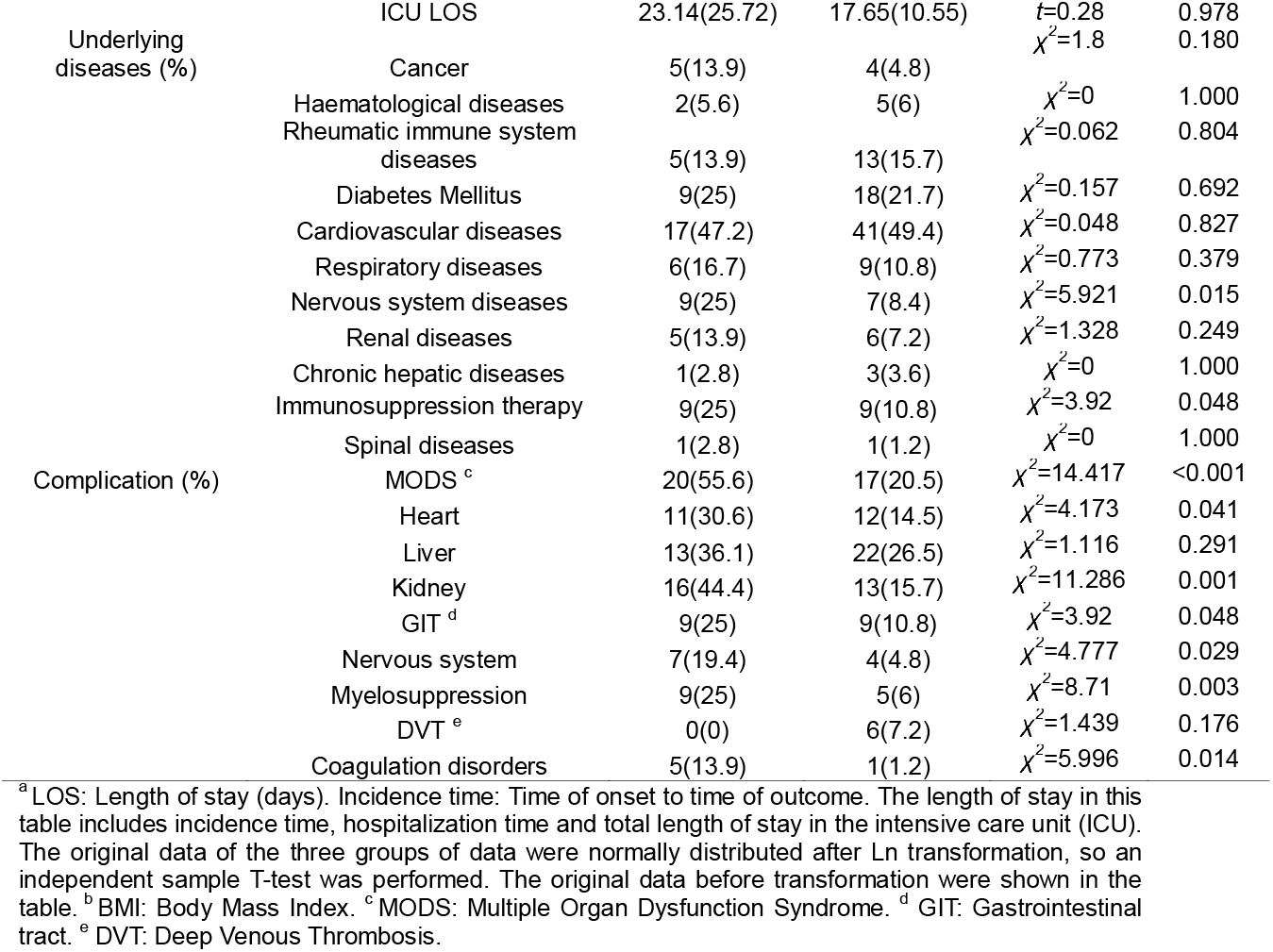
Baseline characteristics.

### Vital signs and laboratory data at admission

Table 2 shows the initial vital signs, laboratory examination results and score scale results for each study group. We recorded the worst vital signs of patients in the first 24 hours before admission to the ICU. The sources of patients included the emergency room and general specialty wards. Some patients from the emergency room were transferred from other community hospitals, and the patients had been given the most basic vital signs support and necessary anti-infective treatment.

**Table2.**
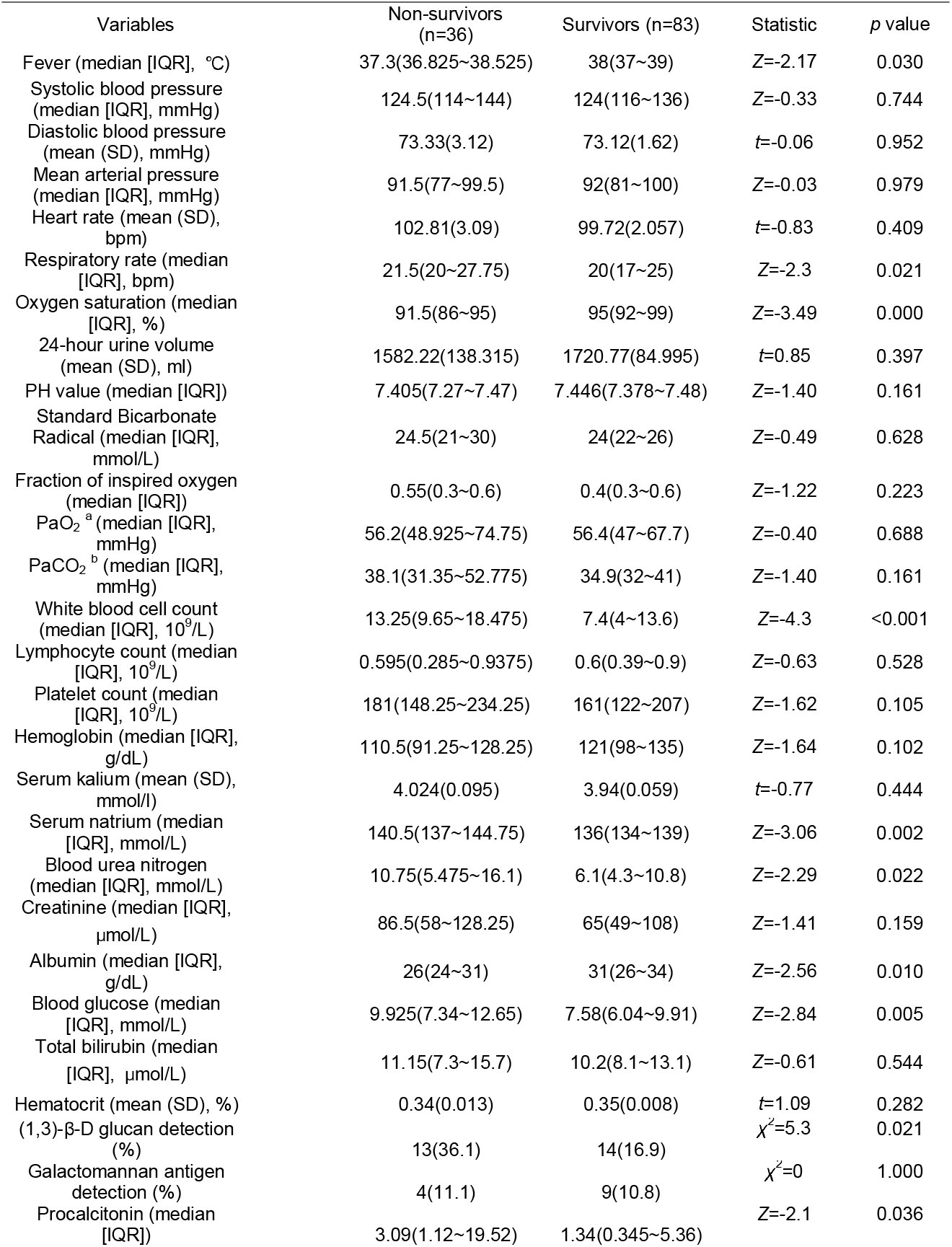
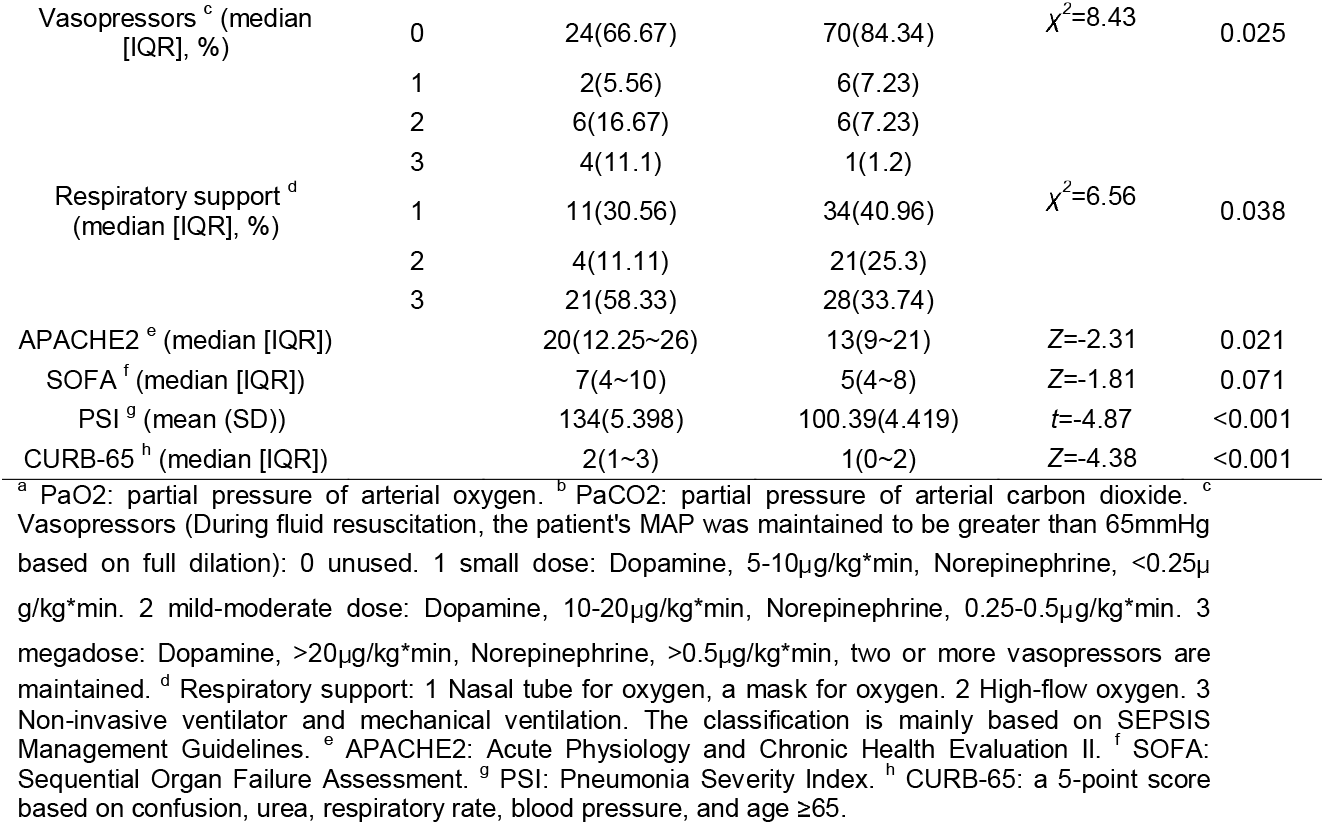
Vital signs and laboratory data at admission.

The initial vital signs we obtain were not necessarily the worst value for the patient throughout SCAP treatment. At the beginning of onset, patients in the non-survivors had already shown worse respiratory performance: faster respiratory rate (*p* = 0.021) and lower oxygen saturation (*p* < 0.001). Non-survivors had lower body temperatures (*p* = 0.03). Urine output during the first 24 hours, acid-base balance and respiratory ventilation were similar in both groups. White blood cell count (*p* < 0.001), serum natrium (*p* = 0.002), blood urea nitrogen (*p* = 0.022) and blood glucose (*p* = 0.005) in the non-survivors were higher, while Albumin (*p* = 0.01) was lower than those in the survivors. In terms of infection markers, the non-survivors had higher procalcitonin (*p* = 0.036) and (1,3)-β-D glucan detection positivity (*p* = 0.021). There was a statistical difference in the use of vasopressors between the two groups at the beginning of disease onset (*p* = 0.025), and the use of moderate dose and megadose Vasopressors accounted for a larger proportion in the non-survivors. And patients requiring ventilator support accounted for a larger proportion in the non-survivors (p = 0.038).

The non-survivors had higher results not only in pneumonia-specific scores: PSI (*p* < 0.001) and CURB-65 (*p* < 0.001), but also in APACHE2 (*p* = 0.021) during the initial score calculation. ROC curves of these scores for mortality showed that AUCs of CURB-65 (OR 0.744 [95%CI, 0.652-0.835], *p* < 0.005), PSI (OR 0.737 [95%CI, 0.65-0.824], *p* < 0.005), and APACHE2 (OR 0.633 [95%CI, 0.522-0.744], *p* < 0.021). Compared with the conventionally used prognosis score for Sepsis, the pneumonia-specific score differed more significantly between the two groups and was more advantageous in guiding prognosis.

### Management during ICU treatment

Table 3 shows the clinical characteristics of each study group during treatment, including fever, respiratory support, and treatment for Septic Shock. For the whole treatment cycle, although the proportion of fever at the beginning of the non-survivors was lower (*p* = 0.002), patients had a longer duration of fever throughout the disease progression (*p* < 0.001). In addition, the time of peak temperature (higher than 39°C) in the non-survivors was also longer (*p* = 0.047). In terms of respiratory support, except for the incidence of tracheotomy, the non-survivors were higher than the survivors in the following items: incidence of tracheal intubation (*p* < 0.001), Mechanical Ventilation (*p* < 0.001), intubated on the NTH day after onset (*p* < 0.001), incidence of more than twice endotracheal intubation (*p* = 0.014), use of Bronchofibroscope (*p* < 0.001), use of Sedatives agents (*p* < 0.001), use of Neuromuscular blocking agents (NMBAs) (*p* < 0.001). We found that the proportion of Septic shock occurred in the non-survivors was higher (*p* < 0.001), duration of Vasopressors used (*p* < 0.001) longer than the survivors. There was no difference in the use of Immunoglobulin between the two groups. In terms of corticosteroid use, only in the total dosage of Corticosteroids (*p* = 0.035), the non-survivors were higher than the survivors.

**Table3.**
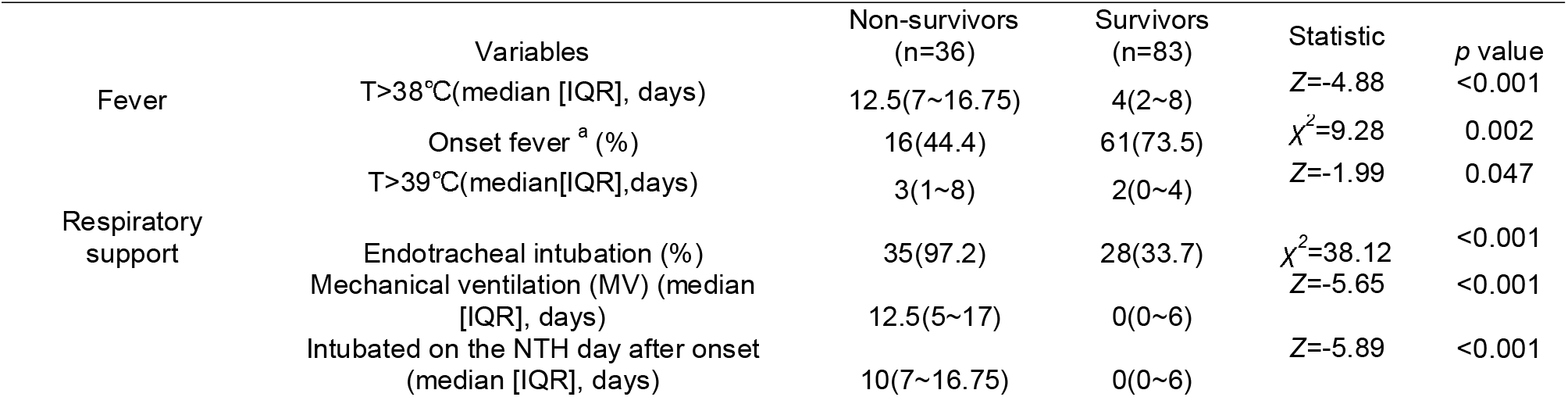
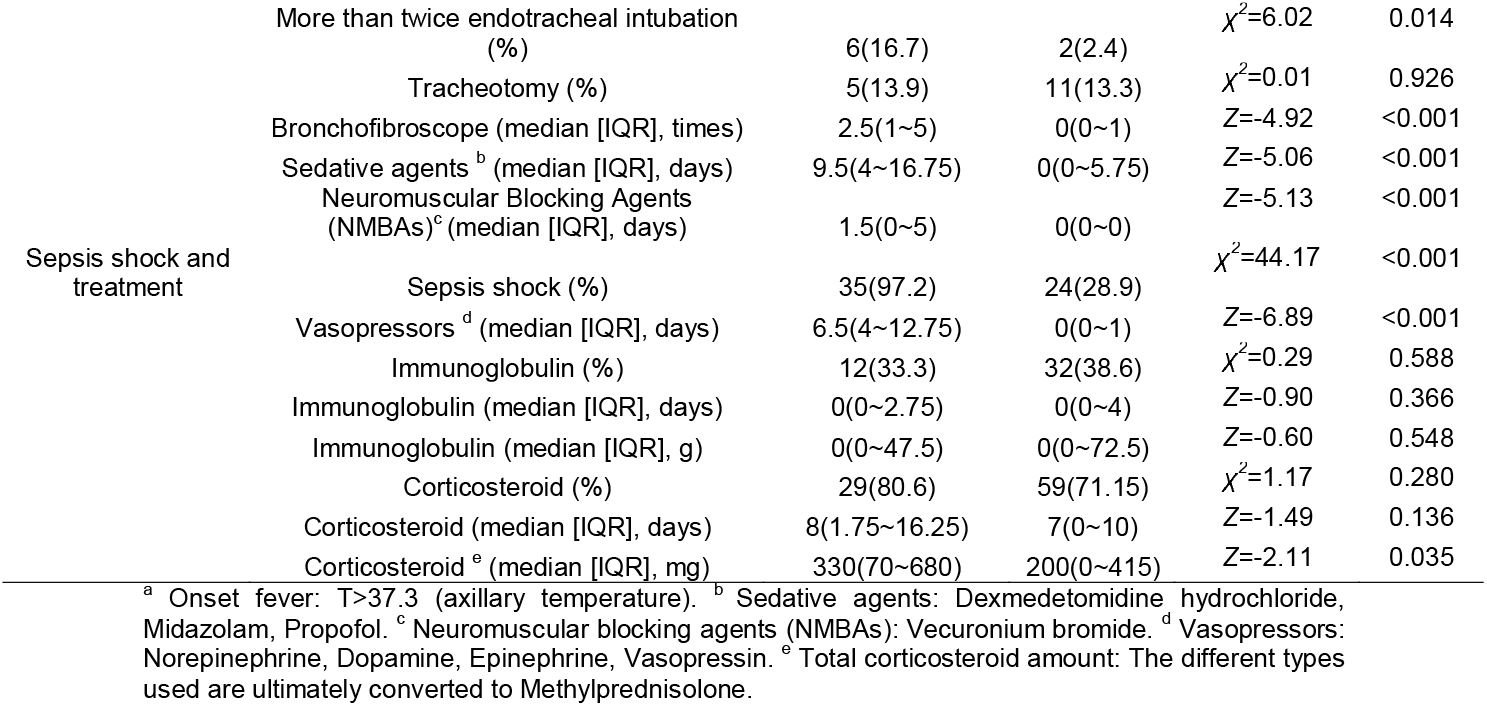
Management during ICU treatment.

### Pathogenic microorganisms and antibiotic use

Table 4 shows pathogenic microorganisms and subsequent antibiotic use in each study group. We counted pathogenic microorganisms throughout the patient’s treatment cycle, including nosocomial secondary infections. Our patients had CAP, most of which were patients with influenza A and B (*p* = 0.002), accounted for 52.1% of the total, and most patients survived treatment. There was no significant difference in atypical pathogen infection between the two groups, including adenovirus, respiratory syncytial virus (RSV), Epstein-Barr virus (EBV), V Herpes Simplex Virus (HSV), Cytomegalovirus (CMV), Pneumocystis carinii pneumonia (PCP), mycoplasma and chlamydia. Patients with SCAP with bacterial onset or secondary bacterial infection had a poor prognosis (*p* = 0.018), patients infected with Gram-negative organisms were more likely to have a poor outcome (*p* = 0.011). The non-survivors had more Blood-borne Infection (BSI) (*p* < 0.001). There was no difference between the two groups in fungal infection, including candida and aspergillus. The two groups were similar in terms of bacterial co-infection (≥2 species of bacteria), and in the number of all pathogenic microorganisms.

**Table4.**
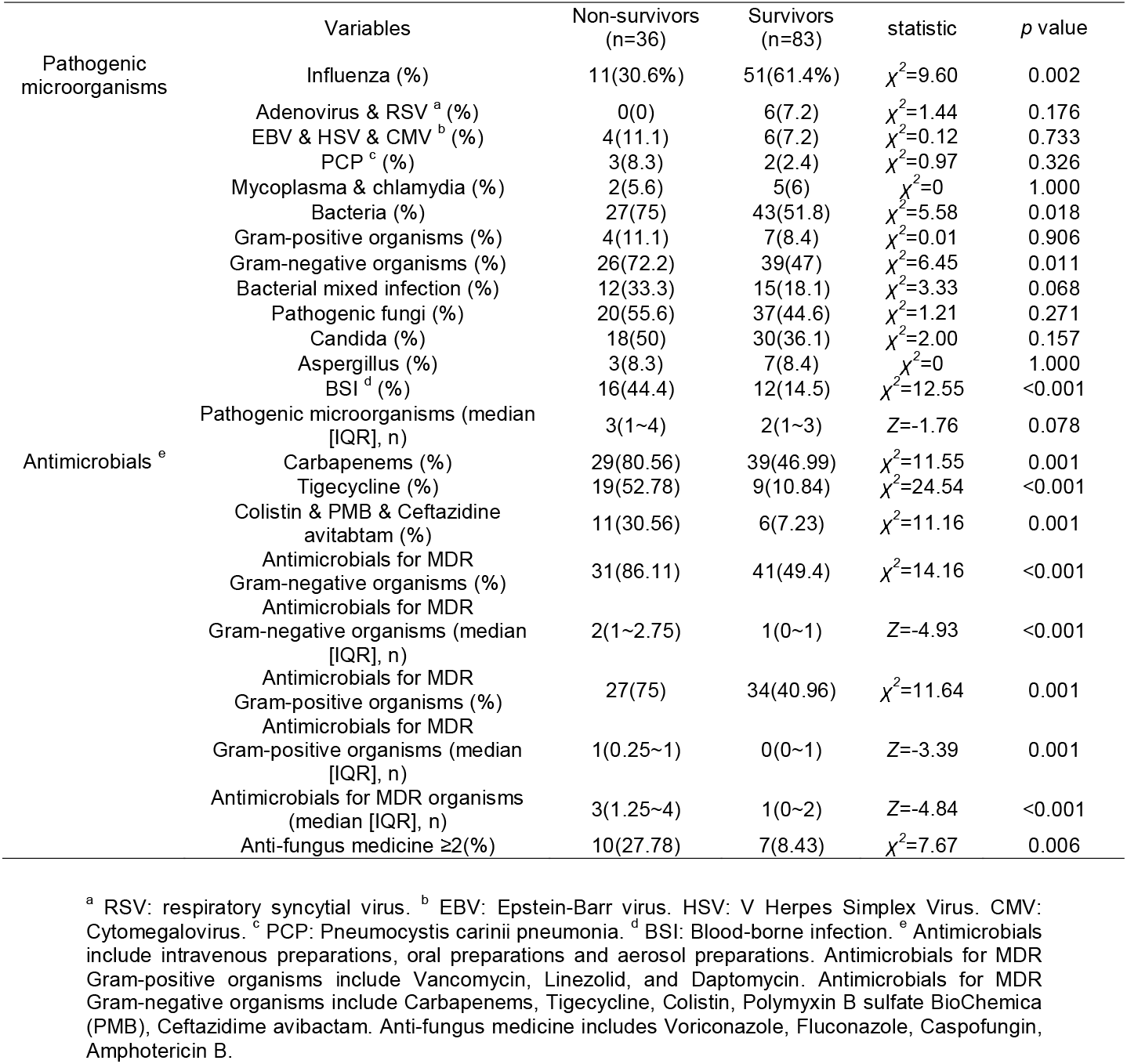
Pathogenic microorganisms and antibiotic use.

In our study, most patients eventually died from refractory pan-drug-resistant infections. Therefore, we mainly evaluated the use of antimicrobials for Multiple Drug Resistance (MDR) in patients. The non-survivors had more use of different types of antimicrobials for MDR, including Antimicrobials for MDR Gram-negative organisms (*p* < 0.001) and Antimicrobials for MDR Gram-positive organisms (*p* = 0.001). Antimicrobials for MDR Gram-negative organisms including: Carbapenems (*p* = 0.001), Tigecycline (*p* < 0.001), Colistin & Polymyxin B sulfate BioChemica (PMB) & Ceftazidime avibactam (*p* = 0.001). In the non-survivors, there were more total antibiotics, including Antimicrobials for MDR Gram-negative organisms (*p* < 0.001) and Antimicrobials for MDR Gram-positive organisms (*p* = 0.001). The non-survivors were more likely to face fungal treatment failure, with a greater proportion of patients with ≥2 species using antifungal agents (*p* = 0.006). We also assessed the total number of Antimicrobials for MDR used in the two groups and found that the non-survivors had a higher number (*p* < 0.001).

### Multivariable Statistical Analyses

The risk factors associated with 180-day mortality in SCAP-Sepsis patients were comorbid with a duration of Vasopressors support, duration of persistent fever, age, numbers of antimicrobials for MDR organisms, CURB-65 score, duration of Neuromuscular Blocking Agents (NMBAs) (OR = 1.234, OR = 1.158, OR = 1.084, OR = 6.484, OR = 3.386, OR = 1.505, respectively). Mechanical Ventilation and Bronchofibroscope were protective factors (table 5). ROC curve for predicting death of each risk factor showed that AUCs were in following: vasopressors 0.866, 95%CI (0.799~0.934), Temperature >38°C 0.778, 95%CI (0.688~0.876), age 0.776, 95%CI (0.678~0.874), antimicrobials for MDR organisms 0.775, 95%CI (0.687~0.864), CURB-65 score 0.748, 95%CI (0.656~0.839), Neuromuscular blocking agents (NMBAs) 0.742, 95%CI (0.638~0.847), *p*< 0.001 (figure 2). The above 6 risk factors had similar effects on 180 days of death prediction of SCAP patients, and all had good predictive value. By calculating the Youden index, we further found that patients with age≥65.5 years, duration of fever ≥9.5 days, number of antimicrobials for MDR organisms ≥2, longer duration of NMBAs and vasopressors, and greater CURB-65 had poorer prognosis.

**Table5.**
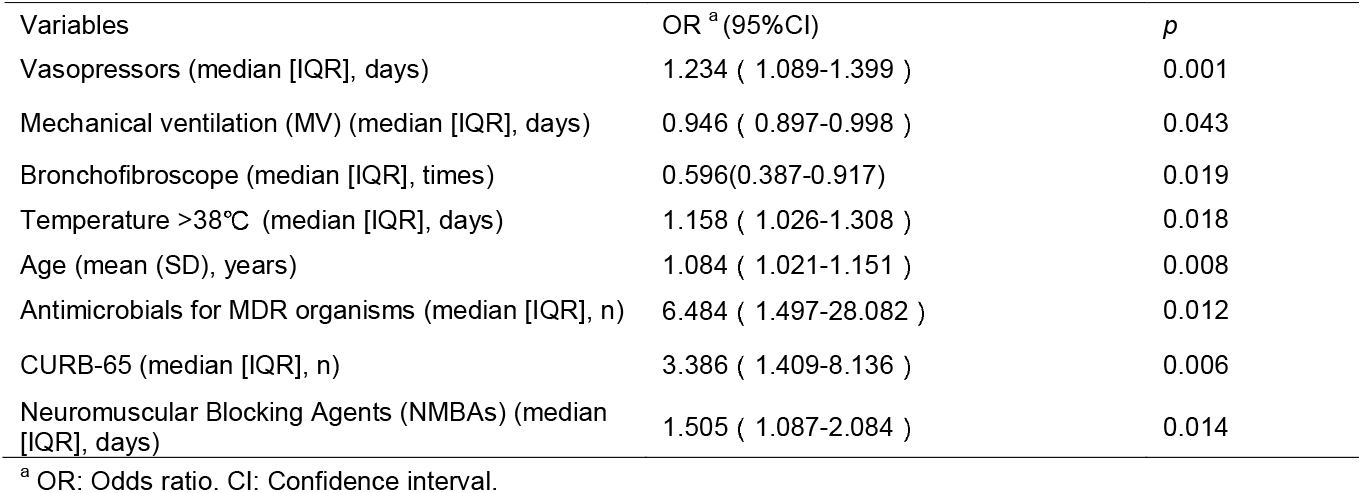
Multivariate analysis of 180-day mortality.

**Figure 2.**
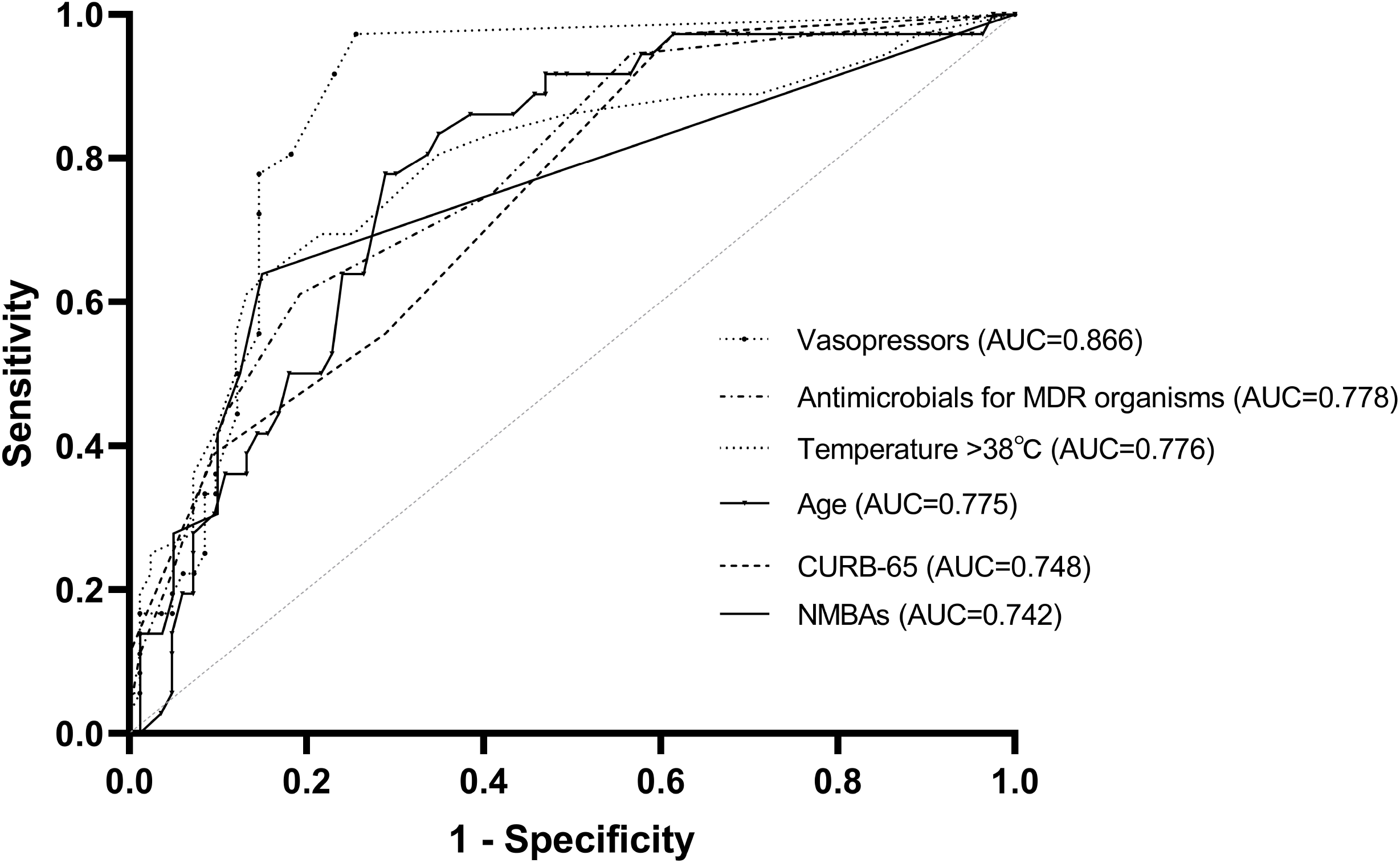
Receiver operating characteristic (ROC) curve of the study population. The grey line is the reference line. NMBAs: Neuromuscular blocking agents.

## DISCUSSION

Our study paid close attention to the patients’ condition throughout the treatment process, which is distinct from previous research focusing on vital signs at a certain time when admitted to the emergency department or ICU. Given the rapid-developing characteristic of Sepsis, the clinical condition at a certain time cannot accurately assess the overall situation and prognosis of the patients, thus an evaluation at a single time can lead clinicians to misjudge a patient’s condition and miss the best time for treatment. The guidelines for Sepsis as well constantly emphasize the significance of dynamic evaluation, while early goal-directed therapy (EGDT) remains controversial, the 2021 International Guidelines for the Management of Sepsis and Shock re-emphasize the adoption of dynamic indicators. [18] Therefore, we evaluated the prognosis of patients by recording the total duration of patients’ outliers in combination with the dynamic situation of patients in the treatment process. This study’s results confirm our hypothesis that a single time point is less valuable than the duration of dynamic monitoring of abnormalities for assessing patients’ outcomes.

This study observed that the temperature of patients before admission to ICU in the non-survivors was lower than that in the survivors, and the proportion of patients with fever at the onset was fewer than that in the survivors. However, the non-survivors had a longer persistent period of fever and the duration of the thermal peak above 39°C was longer than that of the survivors. The deficiency of obvious acute phase responses in patients with Sepsis is associated with high mortality and may reflect the immunosuppressive phase of Sepsis. [10] The non-survivors had similar acid-base balance and pulmonary ventilation function as the survivors at the onset of disease, which may be associated with the corresponding vital sign support, as in Table 2 that the patients in the non-survivors required higher oxygen concentration and higher respiratory support at initial presentation. It is difficult to conclude patients’ prognosis simply from arterial blood gas analysis, which may be interpreted by potential influencing factors including the difficulty of obtaining results before all interventions and the actuality that the worst blood gas indicators can occur at any stage of the disease as a result of patient’s dynamic change of condition.

Our study found the risk factors associated with 180-day mortality in SCAP-SEPSIS patients were comorbid with a duration of Vasopressors support, duration of persistent fever, age, numbers of antimicrobials for MDR organisms, CURB-65 score, duration of NMBAs, which all had a good predictive value. Moreover, the duration of Mechanical Ventilation and frequency of Bronchofibroscope were favourable predictors of outcome.

In this study, the frequency and duration of endotracheal intubation in the non-survivors were significantly higher than those in the survivors, and there were also statistically significant differences in the secondary intubation and intubation time nodes after onset between the two groups. The research results suggest that the persistence time of Mechanical Ventilation and frequency of Bronchofibroscope were protective factors, while the category of antimicrobials for MDR organisms was a risk factor. We supposed that in the secondary ventilator-associated pneumonia (VAP) of SCAP patients, the pulmonary flora disturbance itself might play a more important role than the subsequent infection of microorganisms in the environment with a continuously open airway. No matter how severe the initial lung injury, ARDS patients were prone to suffer secondary lung infection, namely VAP. Bronchial contamination generated by the persistence of endotracheal intubation and Mechanical Ventilation (MV) led to alveolar and systemic defence function impairment, thus pulmonary immune defence and microbiome disorders might play a critical role in ARDS patients. [19] SCAP patients not only presented the contradictory immune status of critical patients but also showed overgrowth of bacteria/fungi and disorder of symbiosis. Moreover, endotracheal intubation, patient’s position, proton pump inhibitors and sedative drug usage all made potential pathogens more favourable for growth, increase in migration, and decrease in elimination. [19–20] Our study found that non-survivors had more disarray of dominant multidrug-resistant bacteria and more categories of antimicrobials for MDR organisms, and that initial treatment failure rate was correlated with prognosis, as well.

Consistent with our conclusion, previous studies had confirmed that the prognosis of disease development was associated with age. [21–22] Menendez et al. found that 11.3% of patients with CAP had more than two organ dysfunctions at diagnosis, which had a higher 30-day mortality rate (12.4% vs. 3.4%) compared with patients without organ dysfunction. [23] Another study discovered that patients with no organ failure had an average mortality rate of 15%, while those with three or more organ failures had a mortality rate of more than 70%.[24] In this study, it was also found that patients who died were more likely to suffer from multiple organ failures than those who survived, but there was no obvious superior choice for the organ prone to failure. It has been suggested that early assessment of organ dysfunction in patients with CAP after admission and even phenotyping analysis of patients with CAP based on the presence of acute respiratory failure or severe Sepsis may facilitate appropriate clinical management and assist in decisions about the location of care. Therefore, it is proposed to assess the severity of the infection and initiate optimal management as early as possible. [23, 25–28]

Our study found that pneumonia-related prognostic scores were superior to sepsis prognostic scores in guiding the outcomes of SCAP patients with Sepsis. It was also mentioned in the latest Sepsis Management Guidelines that qSOFA was not recommended as a single screening tool compared to systemic inflammatory response syndrome (SIRS), National early warning score (NEWS) and modified early warning score (MEWS). We thought that the application of the prognostic score of the disease itself might have more instructive value in the type of sepsis with the definite disease. Similar to our results, Guy Richards et al. found that PSI, CURB-65, and APACHE2 performed the same function in predicting mortality of CAP patients, but CURB-65 and PSI performed better in SCAP patients. [29] A retrospective study of SCAP by Grudzinska FS et al. also supports the use of pneumonia-related prognostic scores for prognostic assessment over general sepsis or early warning scores. They found that CURB-65 was superior to qSOFA or MEWS in the risk stratification of CAP patients. [30] Whereas, Müller M et al. found that qSOFA was superior to CURB-65 in terms of ICU admission prognosis. [31] And Baek MS et al. thought CURB-65 and PSI were not effective in predicting outcomes of elderly patients with pneumonia (older than 80 years). [32]

Our study found that patients in the non-survival group had a lower BMI, as most patients had a BMI within the standard range, and the average BMI in the survival group was even slightly overweight (a BMI of 24 is chosen for the threshold of overweight in China adults). [33–34] In critical patients, we speculated that a certain amount of fat tissue might be protective against disease striking. Previous studies had similarly concluded that a high BMI could reduce mortality in patients with septic shock. [35–37]

This study also found that regardless of the pathogenic microorganism at the onset of the disease, patients with secondary pan-drug-resistant bacterial infection and hematogenous infection had increased difficulty in treatment and poor prognosis. The risk of death increased as the variety and quantity of antimicrobials for MDR organisms increased. If the initial treatment failed, patients with bacteria or fungi infections all faced worse outcomes. Quah J et al. found that respiratory viruses were common isolates of severe community-acquired pneumonia. [38] Virus and bacterial co-infection might increase the risk of death. Harbarth S et al. proposed that appropriate initial antimicrobial therapy was a crucial determinant of survival. [39] Furthermore, Initial appropriate antibiotic treatment outside of ICU for SCAP was also an independent risk factor for prognosis. [40] Resistance to Gram-negative pathogens was a risk factor for inappropriate empiric therapy (IET), and IET could increase the risk of death. In a retrospective cohort study of 175 U.S. hospitals, Zilberberg MD et al. found that Carbapenem-Resistant Enterobacteriaceae (CRE) infection was associated with a four-fold increased risk of receiving IET. [41]

### Limitations and implications for future research

However, our study has some limitations. Firstly, it is a retrospective study with a small sample. Further multicenter studies with large samples are needed. Secondly, statistical data, including respiratory support in later treatment, can be further quantified, such as the pressure value of respiratory support, frequency of prone position ventilation, and frequency of recruitment maneuvers. Thirdly, further research is dedicated to how to recover the normal symbiotic relationship of pulmonary microbial flora, increase bacterial diversity rather than just eliminate dominant species, as well as the relationship between pathogenic microbial disorder and immune response of patients.

## CONCLUSION

It is found that dynamically monitoring the duration of patient abnormalities could help predict patient outcomes. The application of the prognostic score of the disease itself might be more instructive in the type of Sepsis with the definite disease. If the initial treatment failed, both bacteria and fungi infection faced worse outcomes. For SCAP patients with Sepsis, age ≥65.5 years, fever duration ≥9.5 days, number of antimicrobials for MDR organisms ≥2 types, longer NMBAs and Vasopressors use, and higher CURB-65 score tended to predict poorer prognosis.

## Data Availability

All data produced in the present study are available upon reasonable request to the authors

## Ethics approval and consent to participate

The First Affiliated Hospital of Soochow University Ethics Committee (approval No.: 2022-120).

## Consent for publication

Not applicable.

## Availability of data and material

The datasets used and/or analyzed during the current study are available from the corresponding author on reasonable request.

## Authors’ contributions

ZY and CG conceived the study. JJ and ZY designed the study. ZY performed the statistical analysis and drafted the manuscript. JJ was involved in data selection and data collection. CG and ZY contributed substantially to its revision. ZY took responsibility for the manuscript as a whole.

## Open access

This is an open-access article distributed in accordance with the Creative Commons Attribution Non-Commercial (CC BY-NC 4.0) license, which permits others to distribute, remix, adapt, build upon this work non-commercially, and license their derivative works on different terms, provided the original work is properly cited, appropriate credit is given, any changes made indicated, and the use is non-commercial. See: http://creativecommons.org/licenses/by-nc/4.0/.

## REFERENCES

1 Feldman C, Anderson R. Pneumonia as a systemic illness. Curr Opin Pulm Med. 2018;24(3):237–243. doi:10.1097/MCP.0000000000000466

2 Sun Y, Li H, Pei Z, et al. Incidence of community-acquired pneumonia in urban China: A national population-based study. Vaccine. 2020;38(52):8362–8370. doi: 10.1016/j.vaccine.2020.11.004

3 Beutz MA, Abraham E. Community-acquired pneumonia and sepsis. Clin Chest Med. 2005;26(1):19–28. doi: 10.1016/j.ccm.2004.10.015

4 Tsai KS, Grayson MH. Pulmonary defense mechanisms against pneumonia and sepsis. Curr Opin Pulm Med. 2008;14(3):260–265. doi:10.1097/MCP.0b013e3282f76457

5 Schortgen F, Bouadma L, Joly-Guillou ML, Ricard JD, Dreyfuss D, Saumon G. Infectious and inflammatory dissemination are affected by ventilation strategy in rats with unilateral pneumonia. Intensive Care Med. 2004;30(4):693–701. doi:10.1007/s00134-003-2147-7

6 Lin CY, Zhang H, Cheng KC, Slutsky AS. Mechanical ventilation may increase susceptibility to the development of bacteremia. Crit Care Med. 2003;31(5):1429–1434. doi: 10.1097/01.CCM.0000063449.58029.81

7 Pereira JM, Paiva JA, Rello J. Severe sepsis in community-acquired pneumonia--early recognition and treatment. Eur J Intern Med. 2012;23(5):412–419. doi: 10.1016/j.ejim.2012.04.016

8 Rhodes A, Evans LE, Alhazzani W, et al. Surviving Sepsis Campaign: International Guidelines for Management of Sepsis and Septic Shock: 2016. Crit Care Med. 2017;45(3):486–552. doi:10.1097/CCM.0000000000002255

9 Kolditz M, Ewig S, Klapdor B, et al. Community-acquired pneumonia as medical emergency: predictors of early deterioration. Thorax. 2015;70(6):551–558. doi:10.1136/thoraxjnl-2014-206744

10 Hotchkiss RS, Karl IE. The pathophysiology and treatment of sepsis. N Engl J Med. 2003;348(2):138–150. doi:10.1056/NEJMra021333

11 Mandell LA, Wunderink RG, Anzueto A, Bartlett JG, Campbell GD, Dean NC, et al. Infectious Diseases Society of America/American Thoracic Society consensus guidelines on the management of community-acquired pneumonia in adults. Clin Infect Dis. 2007;44(Suppl 2): S27–72. https://doi.org/10.1086/511159.2.

12 Lim WS, Baudouin SV, George RC, Hill AT, Jamieson C, Le Jeune I, et al. BTS guidelines for the management of community acquired pneumonia in adults: update 2009. Thorax. 2009;64(Suppl 3):iii1–55. https://doi.org/10.1136/thx.2009.121434.

13 Fine MJ, Auble TE, Yealy DM, et al. A prediction rule to identify low-risk patients with community-acquired pneumonia. N Engl J Med 1997; 336:243–50.

14 Lim WS, van der Eerden MM, Laing R, et al. Defining community acquired pneumonia severity on presentation to hospital: an international derivation and validation study. Thorax 2003; 58:377–82.

15 Jones AE, Trzeciak S, Kline JA. The Sequential Organ Failure Assessment score for predicting outcome in patients with severe sepsis and evidence of hypoperfusion at the time of emergency department presentation. Crit Care Med 2009; 37: 1649–1654.

16 Knaus WA, Draper EA, Wagner DP, Zimmerman JE. APACHE II: a severity of disease classification system. Crit Care Med. 1985; 13:818–29

17 Cillóniz C, Ewig S, Ferrer M, et al. Community-acquired polymicrobial pneumonia in the intensive care unit: aetiology and prognosis. Crit Care. 2011;15(5):R209. doi:10.1186/cc10444

18 Evans L, Rhodes A, Alhazzani W, et al. Surviving sepsis campaign: international guidelines for management of sepsis and septic shock 2021. Intensive Care Med. 2021;47(11):1181–1247. doi:10.1007/s00134-021-06506-y

19 Luyt CE, Bouadma L, Morris AC, et al. Pulmonary infections complicating ARDS. Intensive Care Med. 2020;46(12):2168–2183. doi:10.1007/s00134-020-06292-z

20 Dickson RP. The microbiome and critical illness. Lancet Respir Med. 2016;4(1):59–72. doi:10.1016/S2213-2600(15)00427-0

21 Marrie TJ, Tyrrell GJ, Majumdar SR, Eurich DT. Effect of Age on the Manifestations and Outcomes of Invasive Pneumococcal Disease in Adults. Am J Med. 2018;131(1): 100.e1–100.e7. doi: 10.1016/j.amjmed.2017.06.039

22 Espinoza R, Silva JRLE, Bergmann A, et al. Factors associated with mortality in severe community-acquired pneumonia: A multicenter cohort study. J Crit Care. 2019; 50:82–86. doi: 10.1016/j.jcrc.2018.11.024

23 Menéndez R, Montull B, Reyes S, et al. Pneumonia presenting with organ dysfunctions: Causative microorganisms, host factors and outcome. J Infect. 2016;73(5):419–426. doi: 10.1016/j.jinf.2016.08.001

24 Martin GS, Mannino DM, Eaton S, Moss M. The epidemiology of sepsis in the United States from 1979 through 2000. N Engl J Med. 2003;348(16):1546–1554. doi:10.1056/NEJMoa022139

25 Aliberti S, Brambilla AM, Chalmers JD, et al. Phenotyping community-acquired pneumonia according to the presence of acute respiratory failure and severe sepsis. Respir Res. 2014;15(1):27. Published 2014 Mar 4. doi:10.1186/1465-9921-15-27

26 Montull B, Menéndez R, Torres A, et al. Predictors of Severe Sepsis among Patients Hospitalized for Community-Acquired Pneumonia. PLoS One. 2016;11(1):e0145929. Published 2016 Jan 4. doi: 10.1371/journal.pone.0145929

27 Welte T. Risk factors and severity scores in hospitalized patients with community-acquired pneumonia: prediction of severity and mortality. Eur J Clin Microbiol Infect Dis. 2012;31(1):33–47. doi:10.1007/s10096-011-1272-4

28 Kolditz M, Braeken D, Ewig S, Rohde G. Severity Assessment and the Immediate and Long-Term Prognosis in Community-Acquired Pneumonia. Semin Respir Crit Care Med. 2016;37(6):886–896. doi:10.1055/s-0036-1592127

29 Richards G, Levy H, Laterre PF, et al. CURB-65, PSI, and APACHE II to assess mortality risk in patients with severe sepsis and community acquired pneumonia in PROWESS. J Intensive Care Med. 2011;26(1):34–40. doi:10.1177/0885066610383949

30 Grudzinska FS, Aldridge K, Hughes S, et al. Early identification of severe community-acquired pneumonia: a retrospective observational study. BMJ Open Respir Res. 2019;6(1): e000438. Published 2019 Jun 5. doi:10.1136/bmjresp-2019-000438

31 Müller M, Guignard V, Schefold JC, Leichtle AB, Exadaktylos AK, Pfortmueller CA. Utility of quick sepsis-related organ failure assessment (qSOFA) to predict outcome in patients with pneumonia. PLoS One. 2017;12(12):e0188913. Published 2017 Dec 21. doi: 10.1371/journal.pone.0188913

32 Baek MS, Park S, Choi JH, Kim CH, Hyun IG. Mortality and Prognostic Prediction in Very Elderly Patients With Severe Pneumonia. J Intensive Care Med. 2020;35(12):1405–1410. doi:10.1177/0885066619826045

33 Zhou B. Cooperative Meta-Analysis Group Of China Obesity Task Force. Zhonghua Liu Xing Bing Xue Za Zhi. 2002;23(1):5–10.

34 Obesity: preventing and managing the global epidemic. Report of a WHO consultation. World Health Organ Tech Rep Ser. 2000;894: i–253.

35 Wurzinger B, Dünser MW, Wohlmuth C, et al. The association between body-mass index and patient outcome in septic shock: a retrospective cohort study. Wien Klin Wochenschr. 2010;122(1-2):31–36. doi:10.1007/s00508-009-1241-4

36 Sato T, Kudo D, Kushimoto S, et al. Associations between low body mass index and mortality in patients with sepsis: A retrospective analysis of a cohort study in Japan. PLoS One. 2021;16(6):e0252955. Published 2021 Jun 8. doi: 10.1371/journal.pone.0252955

37 Gao Q, Cheng Y, Li Z, et al. Association Between Nutritional Risk Screening Score and Prognosis of Patients with Sepsis. Infect Drug Resist. 2021; 14:3817–3825. Published 2021 Sep 17. doi:10.2147/IDR.S321385

38 Quah J, Jiang B, Tan PC, Siau C, Tan TY. Impact of microbial Aetiology on mortality in severe community-acquired pneumonia. BMC Infect Dis. 2018;18(1):451. Published 2018 Sep 4. doi:10.1186/s12879-018-3366-4

39 Harbarth S, Garbino J, Pugin J, Romand JA, Lew D, Pittet D. Inappropriate initial antimicrobial therapy and its effect on survival in a clinical trial of immunomodulating therapy for severe sepsis. Am J Med. 2003;115(7):529–535. doi: 10.1016/j.amjmed.2003.07.005

40 Wongsurakiat P, Chitwarakorn N. Severe community-acquired pneumonia in general medical wards: outcomes and impact of initial antibiotic selection. BMC Pulm Med. 2019;19(1):179. Published 2019 Oct 16. doi:10.1186/s12890-019-0944-1

41 Zilberberg MD, Nathanson BH, Sulham K, Fan W, Shorr AF. 30-day readmission, antibiotics costs and costs of delay to adequate treatment of Enterobacteriaceae UTI, pneumonia, and sepsis: a retrospective cohort study. Antimicrob Resist Infect Control. 2017; 6:124. Published 2017 Dec 6. doi:10.1186/s13756-017-0286-9

